# A simplification of the Kaiser Permanente inpatient risk adjustment methodology accurately predicted in-hospital mortality: A retrospective cohort study

**DOI:** 10.1101/2023.01.06.23284273

**Authors:** Surain B Roberts, Michael Colacci, Fahad Razak, Amol A Verma

## Abstract

**Objective:** We simplified and evaluated the Kaiser Permanente inpatient risk adjustment methodology (KP method) to predict in-hospital mortality, using open-source tools to measure comorbidity and diagnosis groups, and removing troponin, which is difficult to standardize across clinical assays.

**Study Design and Setting:** Retrospective cohort study of adult general medical inpatients at 7 hospitals in Ontario, Canada.

**Results:** In 206,155 unique hospitalizations with 6.9% in-hospital mortality, the simplified KP method accurately predicted the risk of mortality. Bias-corrected c-statistics were 0.874 (95%CI 0.872-0.877) with troponin and 0.873 (95%CI 0.871-0.876) without troponin, and calibration was excellent for both approaches. Discrimination and calibration were similar with and without troponin for patients with heart failure and acute myocardial infarction. The Laboratory-based Acute Physiology Score (LAPS, a component of the KP method) predicted inpatient mortality on its own with and without troponin with bias-corrected c-statistics of 0.687 (95%CI 0.682-0.692) and 0.680 (95%CI 0.675-0.685), respectively. LAPS was well calibrated, except at very high scores.

**Conclusion:** A simplification of the KP method accurately predicted in-hospital mortality risk in an external general medicine cohort. Without troponin, and using common open-source tools, the KP method can be implemented for risk adjustment in a wider range of settings.

## 1 Introduction

The Kaiser Permanente inpatient risk adjustment methodology (KP method) is a well-validated and widely-used method to predict inpatient mortality using routinely collected administrative and laboratory data [1,2]. This method can be used to estimate baseline probability of in-hospital mortality for various applications including quality assessment of different healthcare institutions [3–6]. The KP method has been validated in an external population and demonstrated strong performance [7].

One of the key inputs to the KP method is the laboratory-based acute physiology score (LAPS), which was originally based on 13 laboratory tests, and has since been updated to LAPS2, which added lactate and vital signs [2]. LAPS and LAPS2 include troponin and were originally derived using a specific troponin I assay. This is problematic for implementing the KP method in other settings, because there are numerous troponin I assays and a separate troponin T assay, with varying reference ranges. Linear correlation between these assays does allow some standardization of troponin results, enabling the implementation of LAPS and LAPS2 [8–10]. However, new high-sensitivity cardiac troponin assays were introduced in 2010 and have replaced the older troponin assays in many healthcare institutions [11]. High sensitivity cardiac troponin cannot be harmonized to older troponin assays due to their nonlinear relationship, which makes it impossible to calculate LAPS or LAPS2 as presently formulated [11]. It is not known whether the LAPS and KP method retain predictive ability without the inclusion of troponin. Further, two other aspects of the KP method, the comorbidity points score (COPS) and the grouping of clinical conditions, rely on locally-developed groupings using ICD-9-CM codes, which are not easily implemented in international contexts.

The objective of this study was to evaluate a simplification of the KP method to predict in-hospital mortality in a heterogeneous population of hospitalized medical patients. We replaced COPS and diagnosis groupings with open-source tools that can be easily implemented and we assessed model performance with and without troponin as an input to the LAPS. Furthermore, we investigated whether LAPS accurately predicts the probability of inpatient mortality on its own, to determine its usefulness as an easily-interpreted measure of patient acuity.

## 2 METHODS

### 2.1 Data Source

We conducted a retrospective cohort study using data from 5 academic and 2 community-based teaching hospitals in Ontario, Canada, that are part of GEMINI [12]. GEMINI is a hospital research collaborative that collects administrative and clinical data from hospital information systems with 98-100% accuracy of selected data elements compared to manual chart review [13].

### 2.2 Study population

We included adults 18 years or older who were admitted to or discharged from general medicine between April 1 2010 and December 31 2020. General medicine hospitalizations account for approximately 40% of all emergency admissions at study hospitals [12] and represent a markedly heterogeneous population with no single condition representing more than 5.1% of all hospitalizations [14]. We performed our analysis in a general medicine cohort because it represents a diverse array of conditions and permits a direct assessment of the LAPS score because nearly all patients have pre-admission laboratory measurements from the emergency department. General medicine includes many hospitalizations for cardiovascular conditions [14] where troponin may have particular prognostic value. We excluded hospitalizations during time periods when high sensitivity cardiac troponin assays were performed, to allow us to compare the predictive accuracy of LAPS with and without conventional troponin assays, as it was originally intended.

### 2.3 Implementing the Kaiser Permanente inpatient risk adjustment methodology

Complete details on the derivation and validation of the original KP method have previously been published [1,7]. The variables included in the original derivation are age, sex, admission urgency (elective or emergent), service (medical or surgical), admission diagnosis, severity of acute illness as measured by the LAPS, and chronic comorbidities as measured by the Comorbidity Point Score (COPS). Hospitalizations are grouped by diagnosis and separate logistic regression models are fit within each diagnosis group, allowing for diagnosis-specific intercepts and for each risk factor to have diagnosis-specific coefficients.

The KP method has been updated to include LAPS2 (including lactate and vital signs), mental status, and end-of-life care directives [2]. At our study hospitals, vital signs and end-of-life care directives were documented in paper format for much of the study period and therefore, we implemented the original KP method with LAPS.

The LAPS is a continuous variable calculated by assigning points based on different laboratory values [1]. The theoretical range of the LAPS is 0 to 256, with higher scores denoting greater mortality risk.

We defined diagnosis groups using the Clinical Classifications Software Refined (CCSR) based on a hospitalization’s most responsible discharge ICD-10-CA discharge diagnosis code (or proxy most responsible code where present) [15]. The CCSR method groups all ICD-10 codes into mutually exclusive clinical categories and has been adapted for use with Canadian ICD-10-CA codes by GEMINI [14,16] and is available as an open source software [17]. We took this approach because the original derivation paper used older ICD-9 codes. Other health systems have used CCS, a precursor to CCSR, to derive diagnosis groups for risk adjustment of inpatient mortality [18]. Diagnosis groups with fewer than 150 deaths were grouped together and separated by observed in-hospital mortality rate (>75th, 50th-75th, <50th percentiles). These ranges were selected to ensure all models had sufficient events for parameter estimation and model convergence.

We used the Charlson comorbidity index score as our comorbidity score and only included emergency department and pre-admission diagnosis codes [19]. We did not include medical vs surgical service as a variable because our cohort is restricted to general medicine. We calculated the LAPS using the same weights as the derivation paper, except we did not impute arterial pH, troponin, or total white blood cell count using a 2-step approach and we treat each admission as distinct (no linking of transfers) [1]. LAPS was calculated using the most extreme laboratory values between emergency department triage and time of admission. Laboratory tests that were not performed were assumed to be normal, consistent with the original derivation and other risk adjustment of inpatient mortality [1,18]. Age was squared and modeled as a restricted cubic spline, sex as nominal, admission urgency as nominal, LAPS as linear, and comorbidity score as linear. Two-way interaction terms were included between age squared, LAPS, and comorbidity score [1,7].

### 2.4 Model validation and assessment

We compared the performance of risk adjustment models with and without troponin as part of the LAPS calculation. Models were fit using the entire sample and performance metrics were calculated using Harrell’s bias correction and 1000 bootstrap iterations [20–22]. Resampling was performed before grouping hospitalizations by diagnosis to allow for the number of patients in each diagnosis group to vary between bootstrap iterations. Model performance metrics focused on both discrimination and calibration and included the c-statistic, Brier score, Nagelkerke’s R2, calibration slope, calibration intercept, and visual depiction of calibration curves. We calculated 95% confidence intervals by subtracting the 2.5th and 97.5th percentile optimism values from the apparent value [23].

Bias-corrected calibration curves were calculated for 100 evenly-spaced values within the range of predicted probabilities. Optimism was calculated as the difference in lowess-smoothed curves for refit models in the bootstrapped and original training data. The mean optimism at each value was subtracted from the apparent calibration to produce bias-corrected calibration curves. We only let 5% of points influence the smoother to ensure that deviations from ideal calibration were easily visible (∼2/3 of points is typical [24–26]). All metrics reported in the text are bias-corrected.

Additionally, we investigated the performance of the KP method with and without troponin in the subgroup of hospitalizations for cardiac conditions (CCSR groups for heart failure and acute myocardial infarction). Troponin is known to be prognostic in these conditions [27,28] and so we expected diagnosis-specific models in cardiac conditions to be most affected by removing troponin. We extracted cardiac-diagnosis-specific models from the greater set of models and evaluated them as described above.

We fit univariate logistic regression models agnostic to diagnosis group using only LAPS as a predictor, with and without troponin, and evaluated these univariate models using the methods described above.

All analyses were performed in R version 4.1.0 using the rms package [29,30]. The lrm() function was used to fit logistic regressions and model validation code was adapted from the rms validate() and calibrate() functions. Plotting code for smoothed calibration curves was adapted from the plot.calibrate.default() function.

## 2 RESULTS

### 3.1 Cohort characteristics

Our overall cohort included 353,489 unique hospitalizations from April 2010 to December 2020. High-sensitivity troponin tests were introduced in April 2011, January 2012, November 2014, February 2015, and November 2019 in the 5 academic hospitals and were not introduced at the 2 community hospitals during our study period. After excluding time periods with high-sensitivity troponin testing, our cohort included 206,155 unique hospitalizations.

Median age was 72 years (25th-75th 57-83), 49.3% were female, mean Charlson comorbidity index score at admission was 1.13 (SD 1.67), 29.6% had a Charlson comorbidity index score ≥ 2, 0.5% of hospitalizations were elective, median LAPS was 17 (25th-75th 6-30) and 6.9% of hospitalizations resulted in death (Table 1). The most common diagnosis groups were heart failure (5.1%), pneumonia (4.9%), urinary tract infections (4.5%), chronic obstructive pulmonary disease and bronchiectasis (4.1%), and cerebral infarction (3.5%). LAPS values were similar with and without troponin (quantiles with troponin: 0, 6, 17, 30, 152; quantiles without troponin: 0, 5, 16, 29, 152) (Table 1). There were 23 diagnosis groups with at least 150 deaths, resulting in 23 logistic regression models (equations in the form of R functions are available in the appendix).

**Table 1:** Cohort Characteristics

|  | **Cohort**<br>**(N=206,155)** |
| --- | --- |
| Age - median [Q1 - Q3] | 72.0 [57.0 - 83.0] |
| Female - n (%) | 101,606 (49.3%) |
| LAPS - median [Q1 – Q3] | 17 [6 - 30] |
| LAPS without troponin - median [Q1 – Q3] | 16 [5 - 29] |
| Charlson comorbidity index score at admission – mean (SD) | 1.13 (1.67) |
| Elective admission - n (%) | 1,054 (0.5%) |
| In-hospital mortality - n (%) | 14,265 (6.9%) |
| *Top 10 CCSR diagnosis groups - n (%)* |  |
| Heart failure | 10,428 (5.1%) |
| Pneumonia (except that caused by tuberculosis) | 10,077 (4.9%) |
| Urinary tract infections | 9,296 (4.5%) |
| Chronic obstructive pulmonary disease and bronchiectasis | 8,482 (4.1%) |
| Cerebral infarction | 7,117 (3.5%) |
| Neurocognitive disorders | 6,837 (3.3%) |
| Septicemia | 4,748 (2.3%) |
| Gastrointestinal hemorrhage | 4,737 (2.3%) |
| Intestinal infection | 4,541 (2.2%) |
| Diabetes mellitus with complication | 4,215 (2.0%) |

Abbreviations: laboratory-based acute physiology score, LAPS; Clinical Classification Software Refined, CCSR.

### 3.2 Performance of the KP method

The KP method was able to accurately estimate the risk of inpatient mortality with and without troponin as an input to the LAPS. Bias-corrected c-statistics were 0.874 (95%CI 0.872-0.877) with troponin and 0.873 (95%CI 0.871-0.876) without troponin, indicating strong discrimination (Table 2). Brier scores demonstrated high accuracy in predicted probabilities and were nearly identical with and without troponin (both 0.050, 95%CI 0.050-0.051). Nagelkerke’s R2 values were also similar, suggesting that troponin did little to improve goodness of fit after accounting for all other risk factors (Table 2). The exclusion of troponin from LAPS did not meaningfully affect model calibration, which was excellent as evidenced by calibration intercepts, slopes, and curves (Table 2 and Figure 1 for full performance results).

**Table 2:** Bias-corrected performance of the Kaiser Permanente inpatient risk adjustment methodology, with and without troponin

|  | With Troponin |  | Without Troponin |  |
| --- | --- | --- | --- | --- |
|  | Apparent | Bias-corrected (95% CI) | Apparent | Bias-corrected (95%CI) |
| C-statistic (ROC) | 0.877 | 0.874 (0.872 – 0.877) | 0.876 | 0.873 (0.870 – 0.876) |
| Brier score | 0.050 | 0.050 (0.050 – 0.051) | 0.050 | 0.050 (0.050 – 0.051) |
| Nagelkerke's R<sup>2</sup> | 0.345 | 0.343 (0.337 – 0.349) | 0.347 | 0.341 (0.335 – 0.347) |
| Intercept | 0.000 | - 0.029 (-0.059 – 0.001) | 0.000 | - 0.030 (-0.060 – 0.001) |
| Slope | 1.000 | 0.984 (0.969 – 0.998) | 1.000 | 0.983 (0.969 – 0.997) |

Apparent metrics are based on models in the training data. Bias-corrected metrics are based on models from 1000 bootstrap iterations.

**Figure 1:**
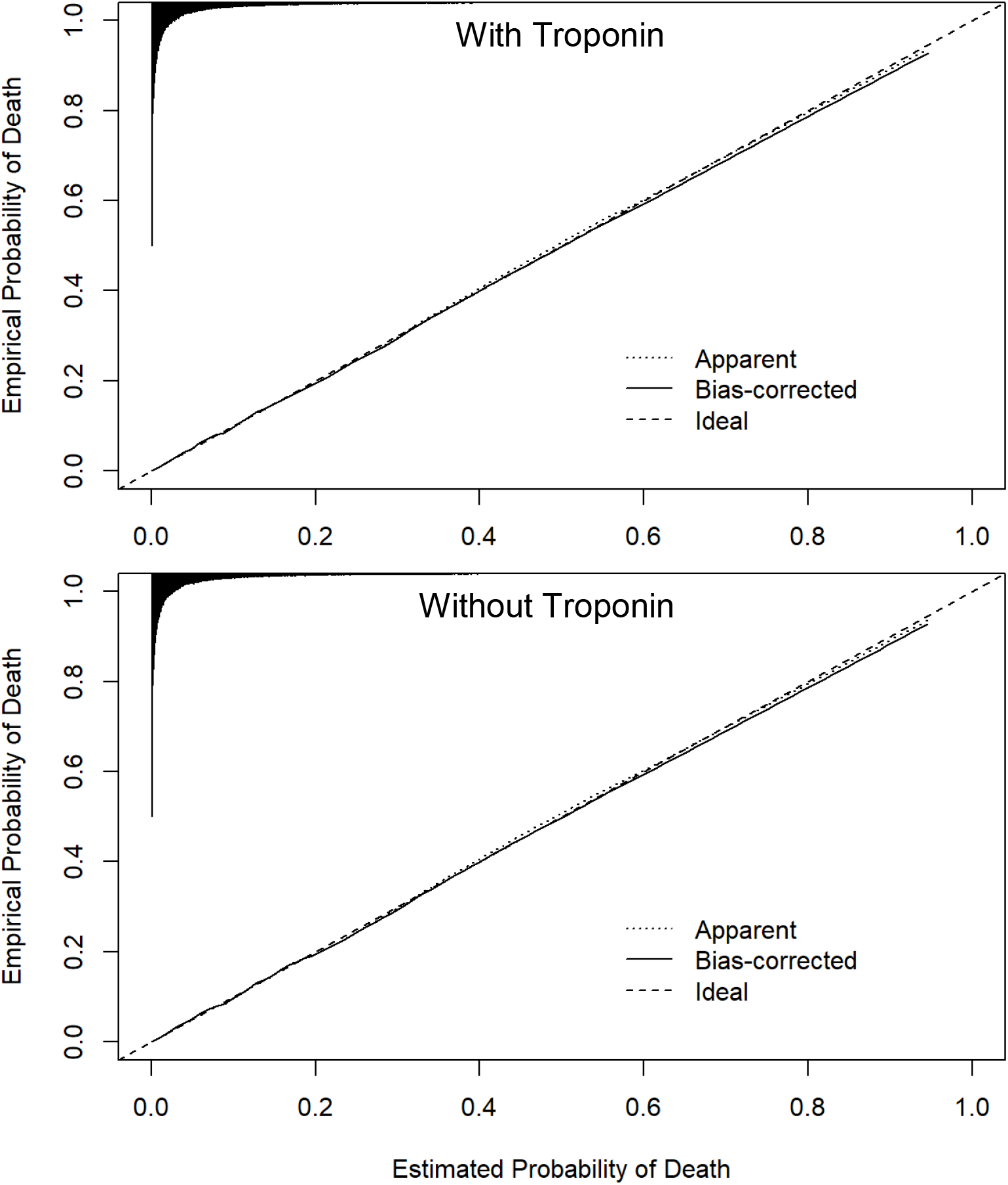
Bias-corrected calibration of the Kaiser Permanente inpatient risk adjustment methodology, with and without troponin Apparent calibration is based on models in the training data. Bias-corrected calibration accounts for optimism through 1000 bootstrap resamples. The histogram on the top denotes the distribution of predicted probabilities and is not scaled to the y-axis.

### 3.3 Performance of the KP method in cardiac conditions

Heart failure and acute myocardial infarction were the two cardiac conditions with sufficient deaths to have their own diagnosis-specific models. The KP method had similar performance with and without troponin in heart failure. For example, the bias-corrected c-statistic with troponin was 0.721 (95%CI 0.703-0.739) and without troponin was 0.717 (95%CI 0.700 - 0.735) (Table 3 and Figure 2 for full performance results). Calibration curves demonstrated that exclusion of troponin led to small improvements in the agreement between high predicted probabilities and observed mortality in the heart failure model, although both models with and without troponin slightly underestimated risk of mortality at high predicted probabilities (Figure 2). The KP method had similar discrimination with and without troponin in acute myocardial infarction (c-statistic with troponin: 0.834, 95%CI 0.810-0.858; without troponin: 0.838, 95%CI 0.815-0.860). Calibration was strong with and without troponin, though without troponin the risk of mortality at high predicted probabilities was very slightly overestimated (Table 4 and Figure 2 for full performance results).

**Table 3:**
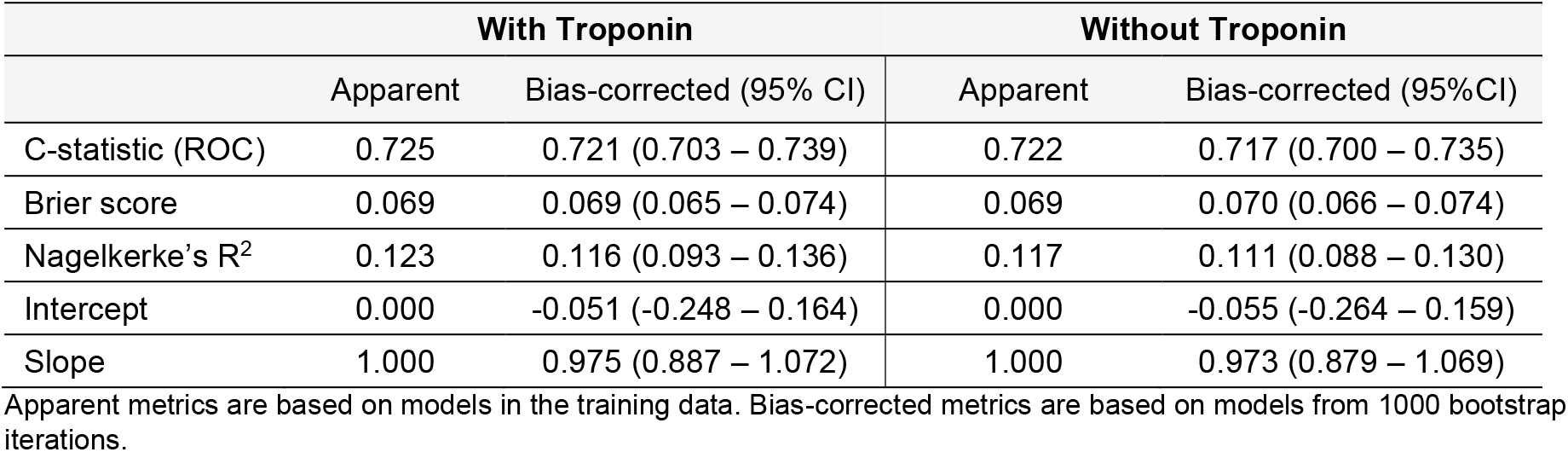
Bias-corrected performance of the heart failure model, with and without troponin

|  | With Troponin |  | Without Troponin |  |
| --- | --- | --- | --- | --- |
|  | Apparent | Bias-corrected (95% CI) | Apparent | Bias-corrected (95%CI) |
| C-statistic (ROC) | 0.725 | 0.721 (0.703 – 0.739) | 0.722 | 0.717 (0.700 – 0.735) |
| Brier score | 0.069 | 0.069 (0.065 – 0.074) | 0.069 | 0.070 (0.066 – 0.074) |
| Nagelkerke's R<sup>2</sup> | 0.123 | 0.116 (0.093 – 0.136) | 0.117 | 0.111 (0.088 – 0.130) |
| Intercept | 0.000 | -0.051 (-0.248 – 0.164) | 0.000 | -0.055 (-0.264 – 0.159) |
| Slope | 1.000 | 0.975 (0.887 – 1.072) | 1.000 | 0.973 (0.879 – 1.069) |

Apparent metrics are based on models in the training data. Bias-corrected metrics are based on models from 1000 bootstrap iterations.

**Table 4:** Bias-corrected performance of the acute myocardial infarction model, with and without troponin

|  | With Troponin |  | Without Troponin |  |
| --- | --- | --- | --- | --- |
|  | Apparent | Bias-corrected (95% CI) | Apparent | Bias-corrected (95%CI) |
| C-statistic (ROC) | 0.841 | 0.834 (0.810 – 0.858) | 0.845 | 0.838 (0.815 – 0.860) |
| Brier score | 0.073 | 0.074 (0.066 – 0.083) | 0.074 | 0.075 (0.067 – 0.083) |
| Nagelkerke's R<sup>2</sup> | 0.293 | 0.273 (0.219 – 0.318) | 0.298 | 0.279 (0.227 – 0.323) |
| Intercept | 0.000 | -0.082 (-0.316 – 0.199) | 0.000 | -0.081 (-0.321 – 0.196) |
| Slope | 1.000 | 0.944 (0.814 – 1.081) | 1.000 | 0.944 (0.814 – 1.081) |

Apparent metrics are based on models in the training data. Bias-corrected metrics are based on models from 1000 bootstrap iterations.

**Figure 2:**
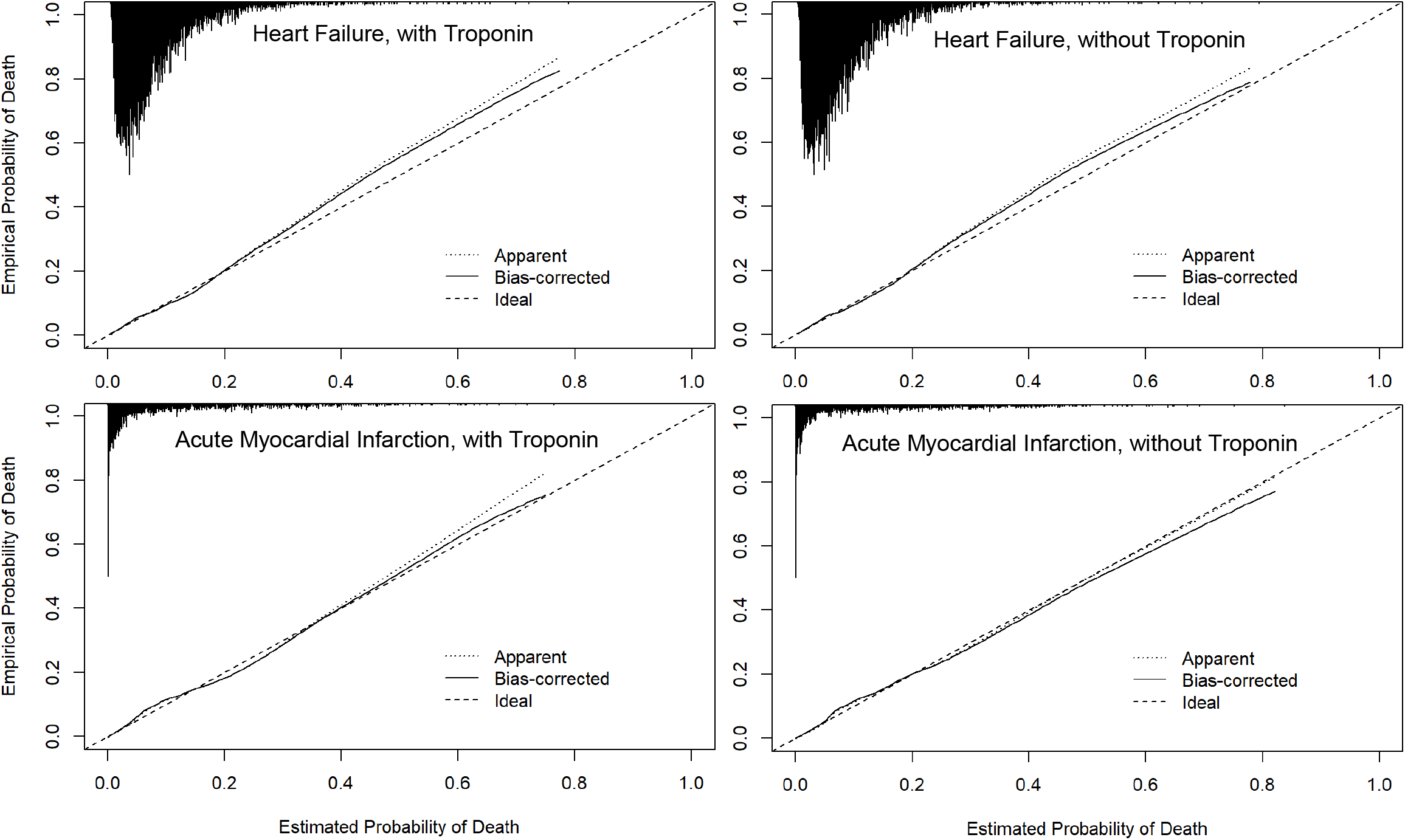
Bias-corrected calibration of the multivariable heart failure and acute myocardial infarction models, with and without troponin Apparent calibration is based on models in the training data. Bias-corrected calibration accounts for optimism through 1000 bootstrap resamples. The histogram on the top denotes the distribution of predicted probabilities and is not scaled to the y-axis.

### 3.4 Univariate analysis of LAPS as a predictor of inpatient mortality

Univariate models including only the LAPS as a predictor showed similar discrimination and calibration with and without troponin in the general medicine cohort (bias-corrected c-statistic with troponin: 0.687 (95%CI 0.682-0.692); without troponin: 0.680 (95%CI 0.675-0.685)(Table 5, Figure 3). Predicted probabilities of death from the LAPS strongly agreed with empirical probabilities when the predicted probabilities were less than approximately 0.3, corresponding to a LAPS of 73. At higher values, the LAPS considerably overestimated the probability of death with and without troponin (Figure 3). Notably, 99% of our cohort had LAPS less than 72 with troponin (and less than 69 without troponin).

**Table 5:** Bias-corrected performance of univariate laboratory-based acute physiology score, with and without troponin

|  | With Troponin |  | Without Troponin |  |
| --- | --- | --- | --- | --- |
|  | Apparent | Bias-corrected (95% CI) | Apparent | Bias-corrected (95%CI) |
| C-statistic (ROC) | 0.687 | 0.687 (0.682 – 0.692) | 0.680 | 0.680 (0.675 – 0.685) |
| Brier score | 0.062 | 0.062 (0.061 – 0.063) | 0.062 | 0.062 (0.061 – 0.063) |
| Nagelkerke's R<sup>2</sup> | 0.081 | 0.081 (0.077 – 0.085) | 0.074 | 0.074 (0.070 – 0.078) |
| Intercept | 0.000 | 0.001 (-0.065 – 0.064) | 0.000 | 0.001 (-0.066 – 0.065) |
| Slope | 1.000 | 1.000 (0.976 – 1.025) | 1.000 | 1.000 (0.974 – 1.026) |

Apparent metrics are based on models in the training data. Bias-corrected metrics are based on models from 1000 bootstrap iterations.

**Figure 3:**
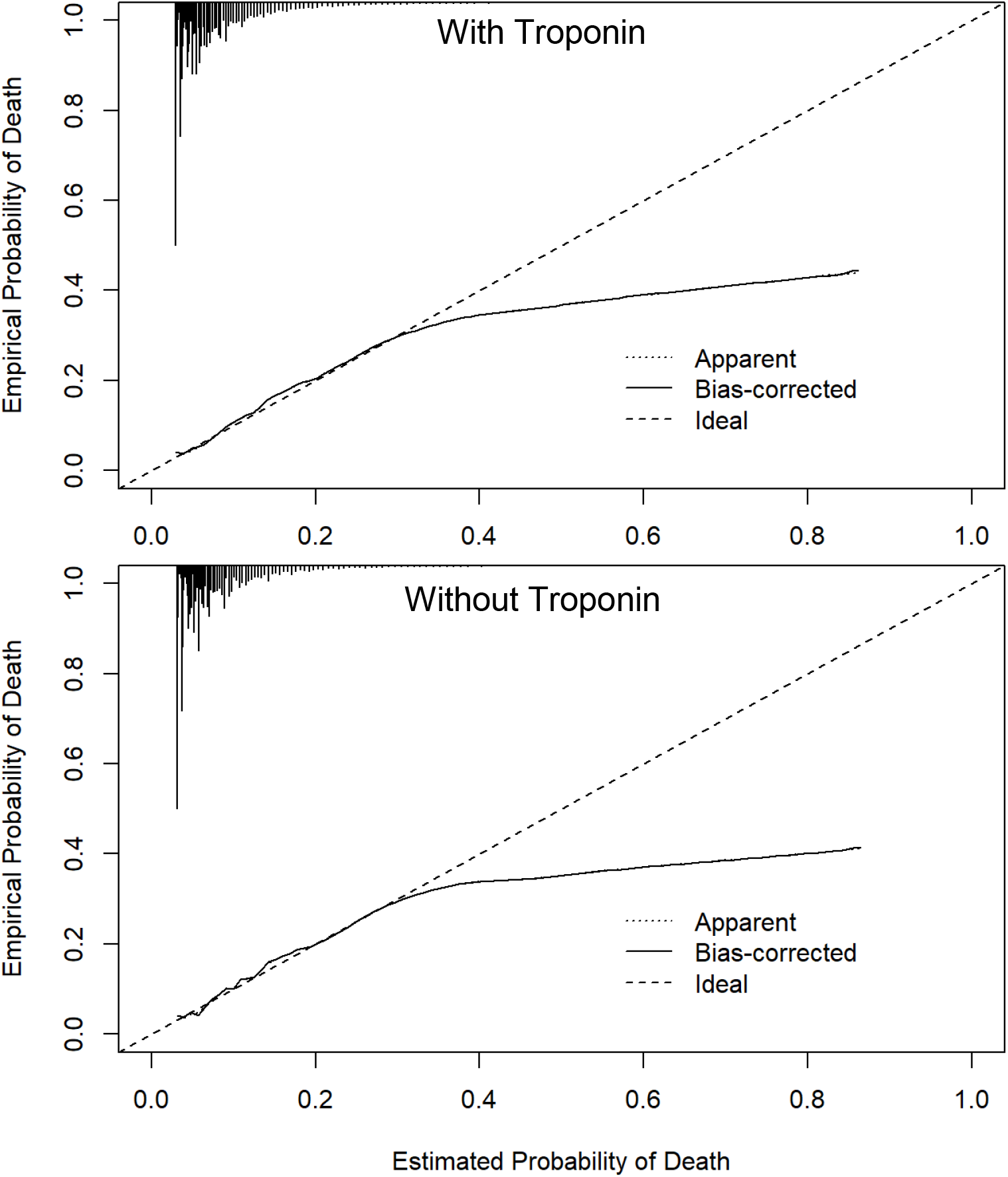
Bias-corrected calibration of univariate model of the laboratory-based acute physiology score, with and without troponin Apparent calibration is based on models in the training data. Bias-corrected calibration accounts for optimism through 1000 bootstrap resamples. The histogram on the top denotes the distribution of predicted probabilities and is not scaled to the y-axis.

Discrimination and calibration of the univariate LAPS models in heart failure and acute myocardial infarction patients are presented in Tables A.1 and A.2 and Figure A.1. Discrimination and calibration were similar with and without troponin in heart failure patients and discrimination was similar in acute myocardial infarction, although calibration was notably worse without troponin in acute myocardial infarction patients.

## 4 DISCUSSION

Among adult general medicine inpatients at 7 hospitals, a simplified version of the Kaiser Permanente inpatient risk adjustment methodology accurately predicted inpatient mortality. Excluding troponin did not meaningfully affect model performance overall or specifically in patients with heart failure or acute myocardial infarction. Importantly, we validate minor changes to the KP risk adjustment system that can greatly simplify its deployment in modern settings. First, we show that it can be used without troponin measurements, which eliminates the need for standardization across conventional troponin assays and enables use in settings that have switched to high-sensitivity troponin measurement. Second, we show the method works well with the Charlson Comorbidity Index score using only diagnosis codes from the current hospitalization rather than the Comorbidity Point Score which requires historical data and ICD-9-CM codes. Finally, we show that the KP method can be deployed using the open-source CCSR software to classify principal diagnoses. Our findings support the external validity of the KP method in a diverse and contemporary cohort and highlight simple adaptations to enhance its adoption.

Many applications in health services research and clinical epidemiology benefit from having a simple, global estimation of inpatient mortality risk, which can be used to describe and compare populations. This study demonstrates that LAPS can serve such a function in heterogeneous inpatient populations. We find that LAPS (with or without troponin) is a good descriptor of inpatient mortality risk for patients with predicted probabilities below 0.30 (LAPS less than 73), which accounts for 99% of adult general medicine patients. However, LAPS on its own is an inferior predictor of inpatient mortality than when it is implemented as part of the broader KP method (c-statistic 0.687 vs 0.874), and LAPS overestimates risk at very high scores.

The transition from conventional troponin T and troponin I to high sensitivity troponin assays has presented challenges for data standardization. The assays measure different enzyme components and are not linearly correlated [11]. Other methods of correlation do not perform well and are not generalizable across diagnosis groups [31]. Our results show that the KP method and LAPS perform very well without troponin. Our findings should not be interpreted to mean that troponin is not a valuable prognostic marker. However, it does not appear to add much predictive value beyond the other variables included in the KP method. The prognostic value of troponin is nicely illustrated in acute myocardial infarction patients, where models with only LAPS suffered a larger decrement in calibration without troponin than multivariable models that included all the risk adjustment inputs.

Similar to a prior external validation study [7], we found strong performance of the KP method. Our cohort (2010-2020) differed substantially from the prior validation cohort (1998-2002). The median LAPS of the previous validation cohort was 0 in comparison to 17 in our study. The most prevalent diagnoses in the previous validation cohort were neurologic disorders (12.9%), arthritis (10.5%) and non-malignant gynecologic disease (8.5%) in comparison to congestive heart failure (5.1%), pneumonia (4.9%) and urinary tract infection (4.5%) in our study. The strong performance of the KP method in both of these external populations, separated by two decades, highlights strong generalizability. Similar to the prior validation, we replaced the COPS (COmorbidity Point Score) with the Charlson Comorbidity Index as a measure of baseline comorbidity. We found that the KP method remains effective when using the Charlson Comorbidity Index score based on pre-admission ICD-10 codes from only that hospital encounter, without historical diagnosis data required to calculate COPS. The original KP method manually grouped diagnoses based on clinical similarity while we use the CCSR classification system, which can be consistently implemented for cohorts defined by ICD-10 codes [16,17].

### 4.1 Limitations

One limitation of this study is that diagnosis groups were categorized according to discharge diagnosis because admitting diagnosis is not reliably available in Canadian administrative health data. We note that prior research has shown that admission and discharge diagnoses are highly correlated within administrative data [32,33]. Though the performance of the KP method was excellent in our study, Escobar et al. have demonstrated that the inclusion of lactate in an updated LAPS2 score, vital signs, and advanced directives further improve model performance [2]. We did not include these additional predictors because they were not consistently available in our dataset and are often missing in large administrative databases [18,34]. Our goal was to validate a more accessible method to encourage adoption by applied stakeholders. Finally, any healthcare institution interested in using the KP method should carefully evaluate the performance of the method in their own data and consider to what extent recalibration might be appropriate.

### 4.2 Conclusion

We validated a simplification of the KP method in a large external population of general medicine patients and demonstrated that it accurately predicts inpatient mortality using common, open-source tools and excluding troponin. These findings support the use of this risk adjustment method in contemporary clinical contexts, particularly if healthcare institutions have transitioned to high sensitivity troponin assays. The KP method remains a rigorous risk adjustment approach for in-hospital mortality.

## Supporting information

Equations as R functions

Dx group mappings

## Data Availability

We are unable to provide unlimited open access to GEMINI data because of data sharing agreements and research ethics board protocols with participating hospitals. However, researchers can request access to GEMINI data through an established process approved by our institutional research ethics boards. Please see full details at: https://www.geminimedicine.ca/access-data

https://www.geminimedicine.gca/access-data

## Author contributions (CRediT taxonomy)

<u>Surain B Roberts:</u> Investigation, Methodology, Formal analysis, Software, Visualization, Writing - original draft, Writing - review & editing

<u>Michael Colacci:</u> Investigation, Visualization, Writing - original draft, Writing - review & editing

<u>Fahad Razak:</u> Resources, Data curation, Supervision, Writing - review & editing

<u>Amol A Verma:</u> Conceptualization, Investigation, Methodology, Resources, Data curation, Supervision, Writing - original draft, Writing - review & editing

## Funding sources

This research did not receive any specific grant from funding agencies in the public, commercial, or not-for-profit sectors. Amol Verma receives salary support as the Temerty Professor of AI Research and Education in Medicine.

## Declaration of Interest

SBR and MC have no disclosures.

Outside of this research, FR and AV are part-time employees of Ontario Health (Provincial Clinical Leads).

## Acknowledgements

We would like to acknowledge the individuals and organizations that have made the data available for this research. The development of the GEMINI data platform has been supported with funding from the Canadian Cancer Society, the Canadian Frailty Network, the Canadian Institutes of Health Research, the Canadian Medical Protective Association, Green Shield Canada Foundation, the Natural Sciences and Engineering Research Council of Canada, Ontario Health, the St. Michael’s Hospital Association Innovation Fund, the University of Toronto Department of Medicine, and in-kind support from partner hospitals and Vector Institute.

## Notes

### Author Declarations

Research ethics board approval was obtained from the University Health Network (Toronto), Sunnybrook Health Sciences Centre (Toronto) and St. Michael's Hospital (Toronto) through the integrated Clinical Trials Ontario platform, with St. Michael's Hospital as the Board of Record (CTO project ID: 1394). Research ethics board approval was also obtained from Trillium Health Partners (Mississauga; REB# 742) and Mount Sinai Hospital (Toronto; REB# 15-0075-C).

