## Appendix for "A simplification of the Kaiser Permanente inpatient risk adjustment methodology accurately predicted in-hospital mortality: A retrospective cohort study"

Table A.1: Bias-corrected performance of the univariate laboratory-based acute physiology score in heart failure patients, with and without troponin

|  | **With Troponin** | | **Without Troponin** | |
| --- | --- | --- | --- | --- |
|  | Apparent | Bias-corrected (95% CI) | Apparent | Bias-corrected (95%CI) |
| C-statistic (ROC) | 0.663 | 0.662 (0.642 – 0.682) | 0.656 | 0.655 (0.635 – 0.676) |
| Brier score | 0.072 | 0.072 (0.067 – 0.076) | 0.072 | 0.072 (0.067 – 0.077) |
| Nagelkerke’s R^2^ | 0.064 | 0.063 (0.046 – 0.078) | 0.059 | 0.057 (0.041 – 0.071) |
| Intercept | 0.000 | 0.002 (-0.256 – 0.307) | 0.000 | 0.002 (-0.271 – 0.314) |
| Slope | 1.000 | 0.999 (0.887 – 1.130) | 1.000 | 0.999 (0.883 – 1.132) |

Apparent metrics are based on models in the training data. Bias-corrected metrics are based on models from 1000 bootstrap iterations.

Table A.2: Bias-corrected performance of the univariate laboratory-based acute physiology score in acute myocardial infarction patients, with and without troponin

|  | **With Troponin** | | **Without Troponin** | |
| --- | --- | --- | --- | --- |
|  | Apparent | Bias-corrected (95% CI) | Apparent | Bias-corrected (95%CI) |
| C-statistic (ROC) | 0.742 | 0.743 (0.707 – 0.779) | 0.757 | 0.757 (0.725 – 0.792) |
| Brier score | 0.081 | 0.081 (0.072 – 0.090) | 0.081 | 0.081 (0.073 – 0.090) |
| Nagelkerke’s R^2^ | 0.145 | 0.143 (0.095 – 0.182) | 0.149 | 0.147 (0.103 – 0.184) |
| Intercept | 0.000 | 0.015 (-0.308 – 0.426) | 0.000 | 0.016 (-0.304 – 0.411) |
| Slope | 1.000 | 1.010 (0.856 – 1.177) | 1.000 | 1.010 (0.868 – 1.172) |

Apparent metrics are based on models in the training data. Bias-corrected metrics are based on models from 1000 bootstrap iterations.

Figure A.1: Bias-corrected calibration of univariate laboratory-based acute physiology score in heart failure and acute myocardial infarction patients, with and without troponin

Heart Failure, with Troponin

Acute Myocardial Infarction, without Troponin

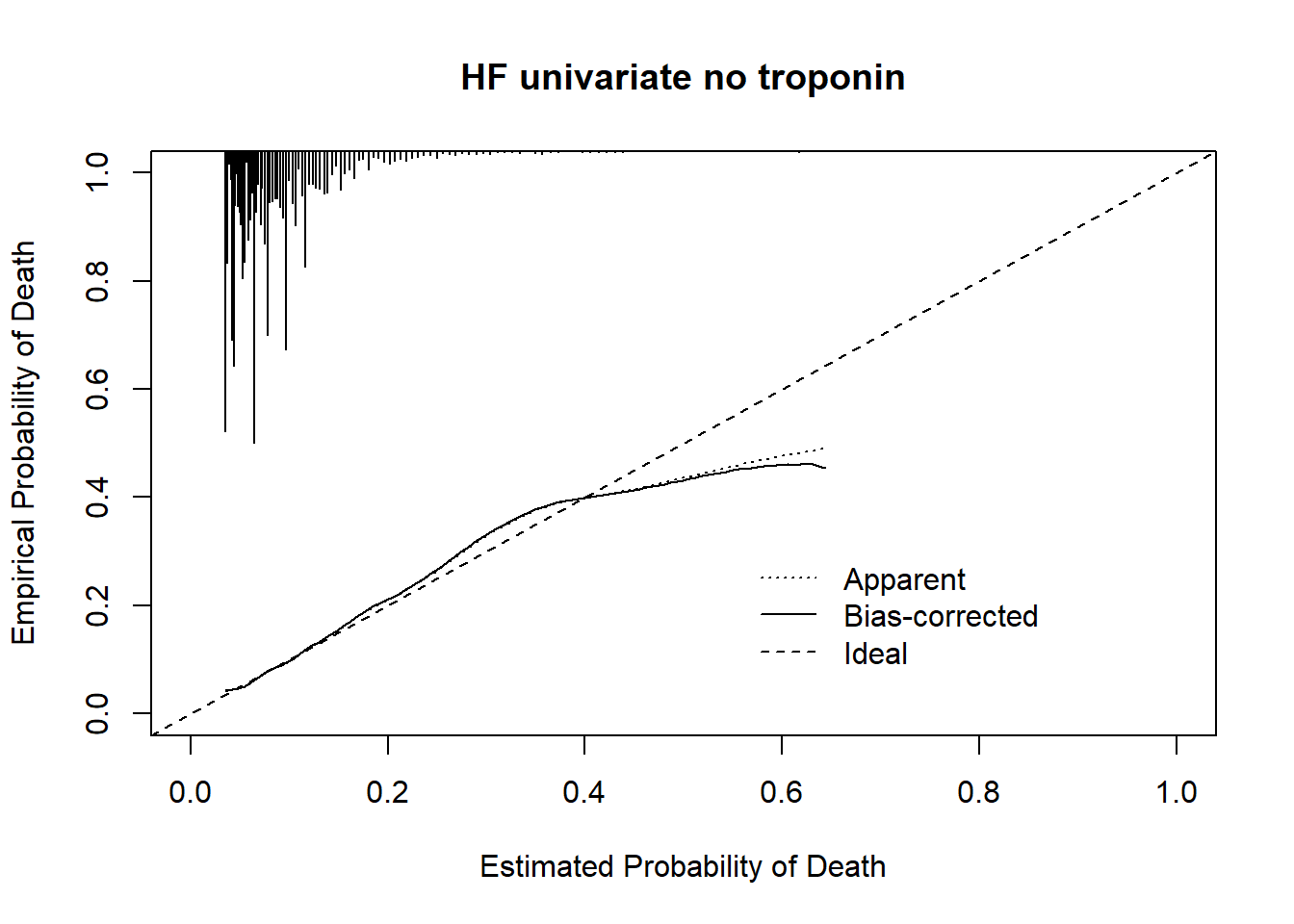

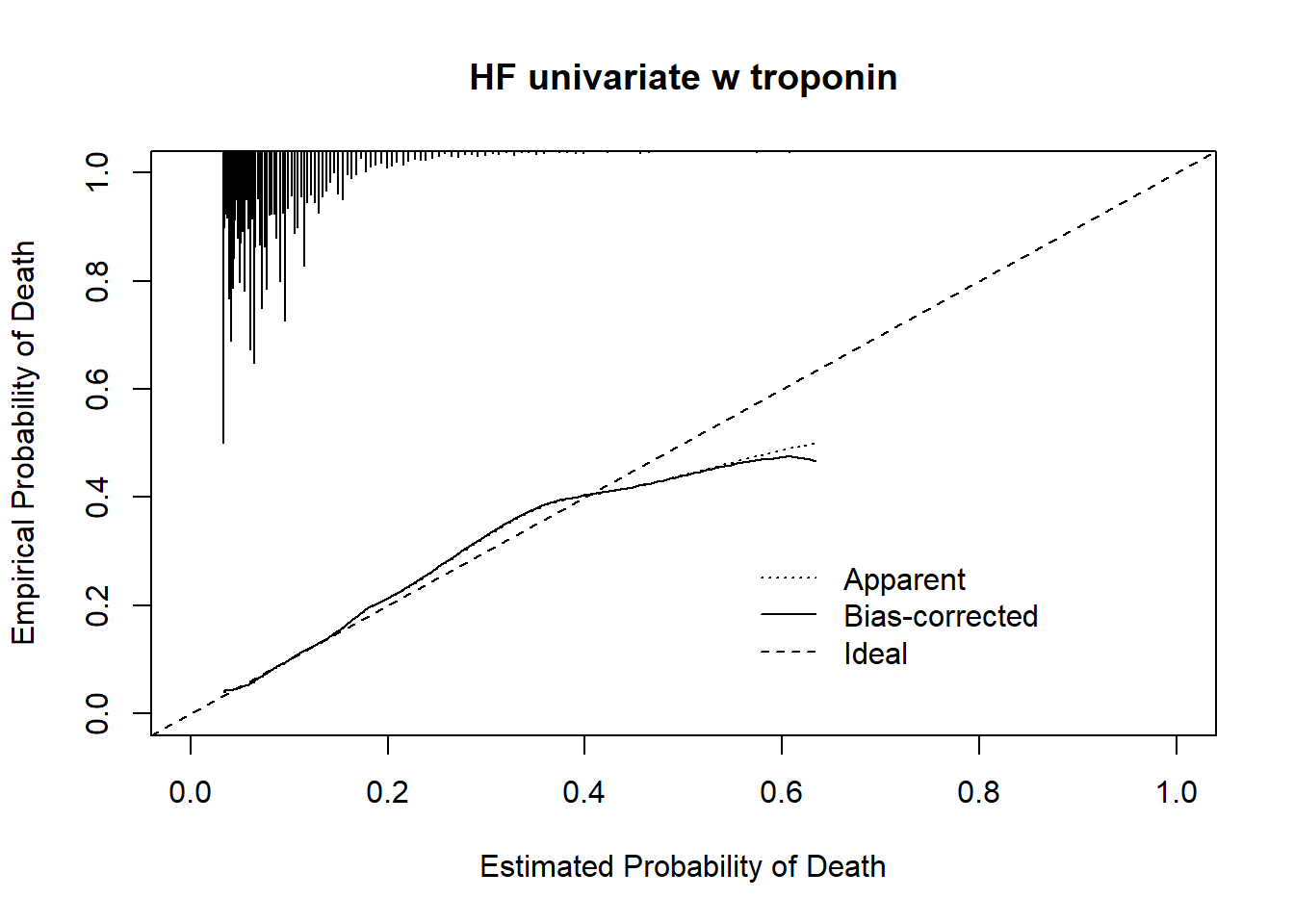

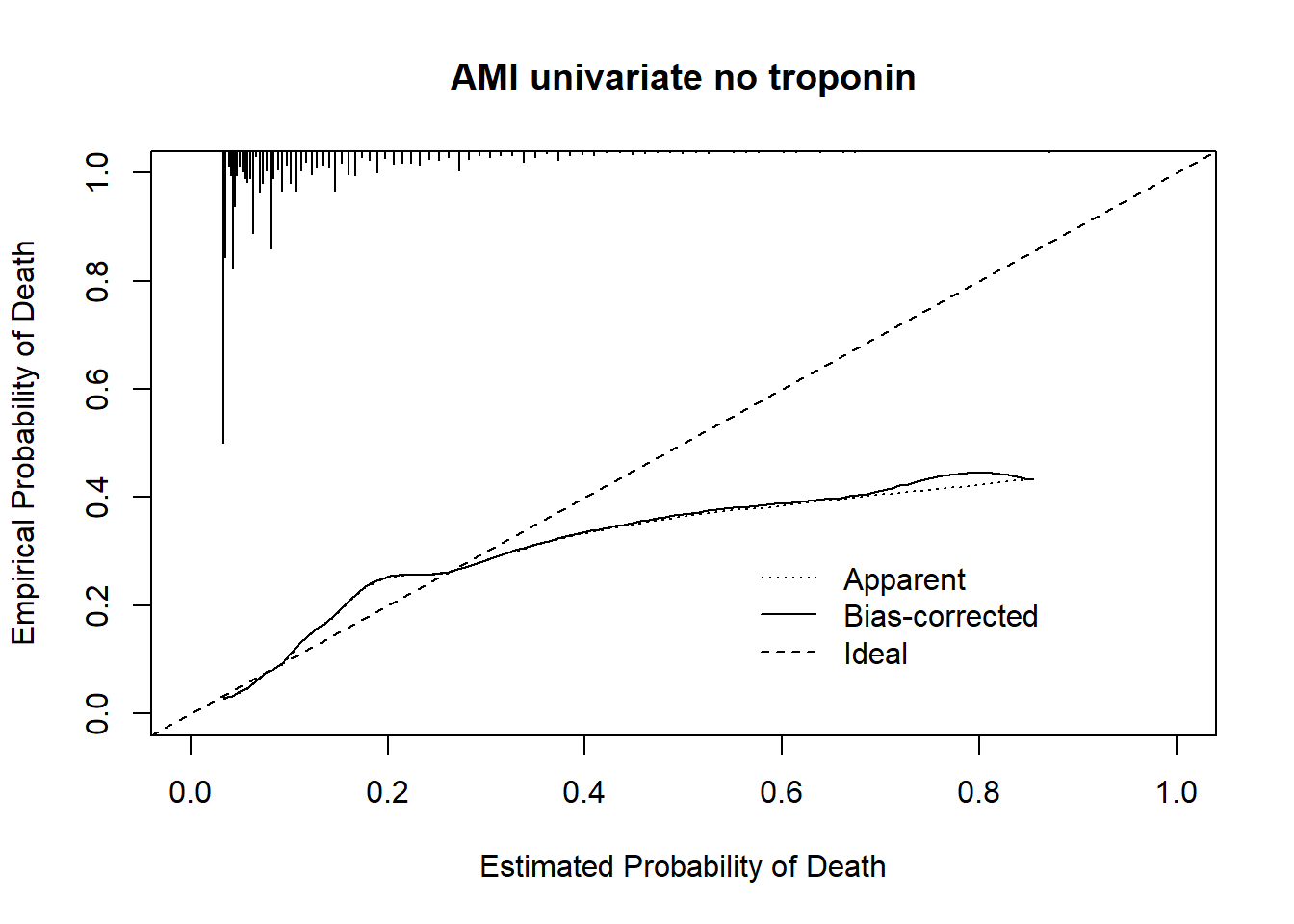

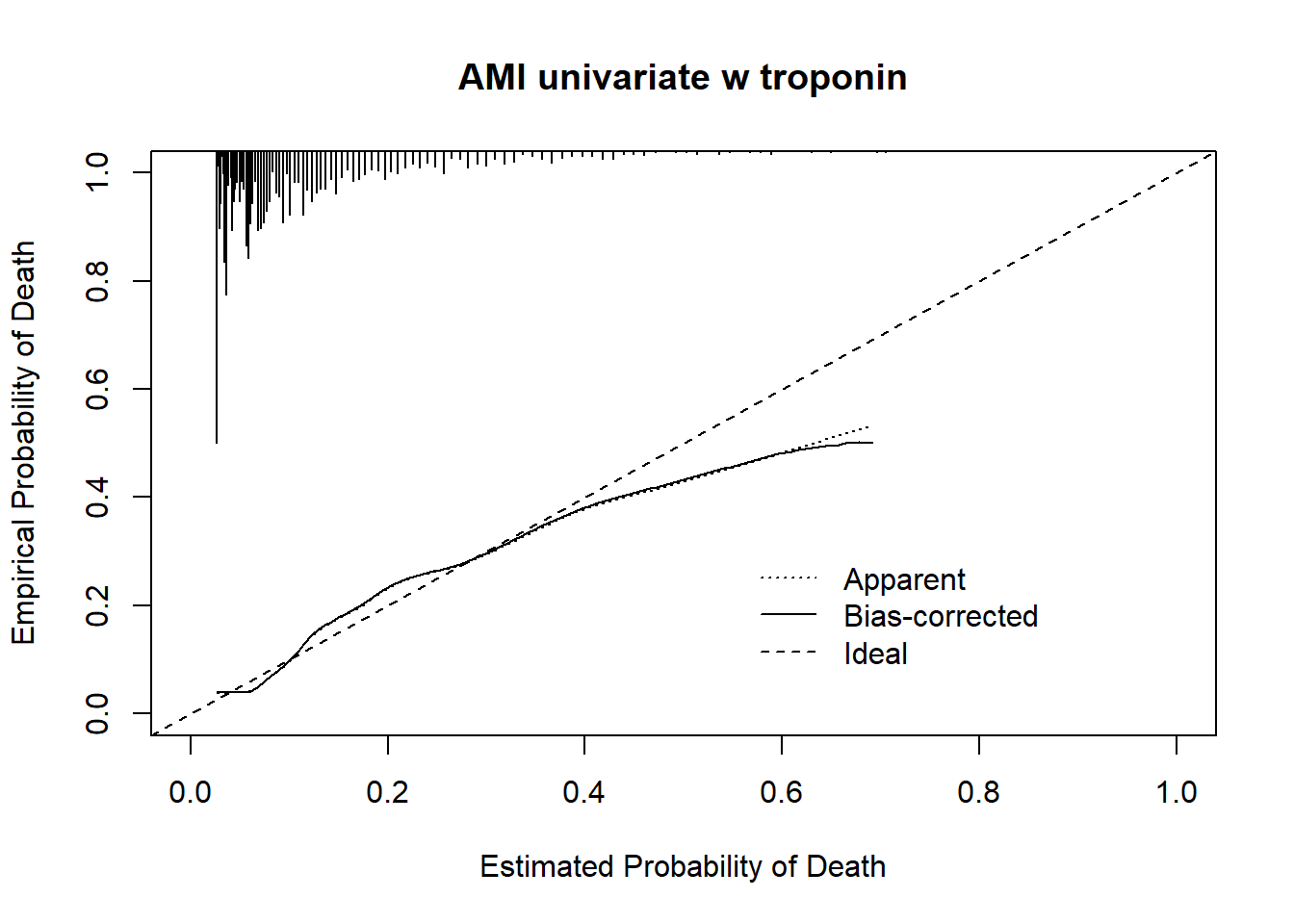

Heart Failure, without Troponin

Acute Myocardial Infarction, with Troponin

Apparent calibration is based on models in the training data. Bias-corrected calibration accounts for optimism through 1000 bootstrap resamples. The histogram on the top denotes the distribution of predicted probabilities and is not scaled to the y-axis.

R Code for Equations:

R functions to generate predicted probabilities based on our implementation of the KP method are available in *equations.tsv.* These functions were generated using Function.transcan() from the Hmisc package (Frank E Harrell Jr (2022). Hmisc: Harrell Miscellaneous. R package version 4.7-0. https://CRAN.R-project.org/package=Hmisc).

The first column named ‘equation’ is the R function itself, and the second column named ‘ccsr_mod’ is the diagnosis group corresponding to the function. Diagnosis groups in your data should be coded using [CCSR](https://www.hcup-us.ahrq.gov/toolssoftware/ccsr/ccs_refined.jsp), which can be implemented using our [open source software](https://github.com/GEMINI-Medicine/gemini-ccsr) in Python.

See *mappings.tsv* to preprocess CCSR categories in your data to match the ‘ccsr_mod’ column in *equations.tsv*. The first column in *mappings.tsv* is ‘ccsr_mod’ and is the diagnosis group corresponding to the functions. The second column is ‘ccsr_desc’ which includes the original un-collapsed CCSR categories.

These functions expect variables to adhere to the following coding:

| **Variable** | **Coding** | **Note** |
| --- | --- | --- |
| gender | F, NF | F = female, NF = not female |
| age2 | Numeric | Age squared |
| admit_category | Elective, Urgent | Surgical patients were not included in this study |
| admit_charlson_derived | Numeric | Charlson comorbidity index score (Quan 2011) |
| laps_nt | Numeric | LAPS score without troponin, using same scoring as the derivation paper |

Our models can be implemented in R as functions using the following code. This code generates a list with each function as a separate element, and one element per diagnosis group. Note that this R code is provided for illustrative purposes only and comes with absolutely no warranty.

\# ---

\# Read in equations

equations <- read.table("equations.tsv", sep = '\t', header = TRUE, quote = "\\")

\# Create a list with each equation as an element

funcs_char <- split(equations, f = ~ ccsr_mod)

#Evaluate the equations as R functions

funcs_char <- lapply(funcs_char, function(x) str2lang(x$equation))

funcs <- lapply(funcs_char, function(x) eval(x))

\# Example prediction for a patient in the heart failure group, on the logit scale

funcs$`Heart failure`(gender = "F",	# female patient

age2 = 75^2,	# 75 years old

admit_category = "Urgent", # Urgent admission

admit_charlson_derived = 4, # Charlson comorbidity index score of 4 at admission

laps_nt = 20)	# LAPS score without troponin of 20 at admission

\# ---
